# Microbiological Assessment of Health-Care Providers in Africa: Systematic Review and Meta-Analysis

**DOI:** 10.1101/2021.10.14.21264931

**Authors:** Abeer B. Idris, Alaa B. Idris, Elfatih A. Hasabo, Marwan M. Badawi, Nazar Beirag

## Abstract

**Background:** Healthcare workers (HCWs) are the safeguards that help prevent illnesses and eliminate them when they occur. This study aimed to scan the related literature and provide pooled data about the level of knowledge/attitude toward infections, vaccination status and infection prevalence among African HCWs to provide better evidence toward specific detailed determination of gaps to strengthen. A total of 11,038 published articles were identified from the search strategy. Among them, 163 articles met our inclusion criteria and passed the quality assessment procedure.

**Results:** The prevalence of HBV was tested for 6,599 African HCWs;6.00% [95% Cl; 3.66, 8.33] were positive. The question Are you fully vaccinated against HBV?” was answered by 12,036 HCWs; 43.22% [95% Cl; 31.22, 55.21] answered yes. The most crucial local factor identified among respondents for the spread of antimicrobial resistance (AMR) was self-antibiotic prescription 42.00 % [18.79, 65.20]. The question “Does the infection prevention and control (IPC) guidelines available in your workplace?” was asked to 1,582 HCWs; 50.95% [95% Cl; 40.22, 61.67] answered yes.

**Conclusion:** This study determined many weaknesses to be addressed for the sake of improving health in Africa. The current pooled data are critically significant to be implemented in planning governmental or NGOs strategies.

## 1. Introduction

Healthcare workers (HCWs) are the safeguards that assist in preventing illnesses and eliminate them when they arise (1). However, the fact that they are a double-edged sword is not to be underestimated (3). When standards go low, safeguards are needed to be saved, and more importantly, they may jeopardize others (4). Several studies indicated that a vast percent of patients (up to 50 %) are admitted to hospitals after suffering from healthcare- associated infections (5).

Moreover, Africa is considered the continent with the lowest Gross Domestic Product (GDP) as most African countries fall within the lower-middle to low- income countries classification, which could be directly altering continuous education for healthcare workers, weak healthcare infrastructure, application of infection control policies in health facilities and vaccination opportunities against vaccine-preventable diseases for both healthcare workers and the community (7)(8). Furthermore, sub-Saharan Africa accounted for 66% of the human immunodeficiency viruses (HIV) cases, 68% of new adult HIV infections, 92% of new infections in children and 72% of all acquired immunodeficiency syndrome (AIDS) - related deaths, according to the Joint United Nations Program on HIV/AIDS (UNAIDS) in 2017, while South Africa is the country with the world’s most considerable prevalence of people living with HIV as 5.6 million (2). On the other hand and according to the World Health Organization (WHO), the highest prevalence of hepatitis B virus (HBV) infection is reported in the Western Pacific Region, and the African Region as 6.2% and 6.1% of the adult population are infected, respectively (9). Furthermore, Sub-Saharan Africa was also identified as having an immense burden of human papillomaviruses (HPV) in the world (24.0%) (10). Regarding tuberculosis infection (TB), WHO in 2015 identified two African countries (Nigeria and South Africa) alongside the other three Asian countries to be responsible for 60% of incident cases (6).

Many studies have identified infection prevalence, vaccination status, knowledge level, and attitudes among African HCWs regarding various diseases. Therefore, the current systematic review aimed to scan the related literature and provide pooled data about the level of knowledge/Attitude toward infections, vaccination status and infection prevalence among African HCWs to provide better evidence toward specific detailed determination of gaps to strengthen.

## 2. Methods

### 2.1 Search strategy

To identify relevant studies, a systematic review of the literature was conducted on July 1, 2020. The review was conducted following the PRISMA (Preferred Reporting Items for Systematic Reviews and Meta-Analyses) Statement (11). A comprehensive search was conducted in PubMed, Embase, Google Scholar, Scopus, Index Copernicus, DOAJ, EBSCO-CINAHL, Cochrane databases without language limits (studies written in French were later excluded). To obtain current situation evidence, only studies published in or after 2010 were included. Furthermore, all studies where the data collection process took place before 2010 were also excluded. The only exception was if the collection process continued months/years earlier than 2010 and ended in 2010 or afterwards as previously described (12). The keywords used in PubMed was as follow:

((specialists[ti] OR technologists[ti] OR technicians[ti] OR nurses[ti] OR clinicians[ti] OR physicians[ti] OR doctors[ti] OR medical[ti] OR laboratory[ti] OR hospital [ti] OR “health personnel’’[ti] OR “health staff’’[ti] OR healthcare[ti] OR Dentists [ti] OR health[ti]) AND (behavior[ti] OR risk[ti] OR awareness[ti] OR Attitude[ti] OR knowledge[ti] OR assessment[ti] OR evaluation[ti] OR Measure[ti] OR vaccination[ti] OR Prophylaxis[ti] OR screening[ti] OR infection[ti] OR Injury[ti] OR survey[ti] OR Practice[ti] OR surveillance[ti] OR prevalence[ti] OR incidence[ti] OR bacteria[tiab] OR fungi[tiab] OR virus[tiab] OR viral[tiab] OR bacterial[tiab] OR fungal[tiab]) AND (africa[Tiab] OR algeria[Tiab] OR angola[Tiab] OR benin[Tiab] OR botswana[Tiab] OR burkinafaso[Tiab] OR burundi[Tiab] OR caboverde[Tiab] OR cameroon[Tiab] OR central african republic[Tiab] OR CAR[Tiab] OR chad[Tiab] OR comoros[Tiab] OR “democratic republic of the congo’’[Tiab] OR “republic of the congo’’[Tiab] OR cote d’ivoire[Tiab] OR djibouti[Tiab] OR egypt[Tiab] OR equatorial guinea[Tiab] OR eritrea[Tiab] OR eswatini[Tiab] OR swaziland[Tiab] OR ethiopia[Tiab] OR gabon[Tiab] OR gambia[Tiab] OR ghana[Tiab] OR guinea[Tiab] OR guinea-bissau[Tiab] OR kenya[Tiab] OR lesotho[Tiab] OR liberia[Tiab] OR libya[Tiab] OR madagascar[Tiab] OR malawi[Tiab] OR mali[Tiab] OR mauritania[Tiab] OR mauritius[Tiab] OR morocco[Tiab] OR mozambique[Tiab] OR namibia[Tiab] OR niger[Tiab] OR nigeria[Tiab] OR rwanda[Tiab] OR sao tome principe[Tiab] OR senegal[Tiab] OR seychelles[Tiab] OR sierra leone[Tiab] OR somalia[Tiab] OR south africa[Tiab] OR south sudan[Tiab] OR sudan[Tiab] OR eswatini[Tiab] OR tanzania[Tiab] OR togo[Tiab] OR tunisia[Tiab] OR uganda[Tiab] OR zambia[Tiab] OR zimbabwe[Tiab])).

Moreover, to optimize our search, hand searches of reference lists of included articles were also performed.

### 2.2 Study selection and data extraction

All authors independently assessed titles and abstracts for eligibility, and any disagreement was resolved through discussion. A copy of the full text was obtained for all research articles that were available and approved in principle to be included. Abstraction of data was under the task separation method; method and result sections in each study were separately abstracted on different occasions to reduce bias. Moreover, data abstraction was conducted without considering the author’s qualifications or expertise (13). When included study gathers groups of health workers and other workers as participants with detailed results for each group; only the health worker participant results will be included, if no detailed results are available, the study will be excluded. If data regarding the period of conduction is missing in a study, the reference list will read and if any cited study is published after 2010, the authors of the current review agreed to predict that the study is conducted after 2010, hence it was considered for inclusion and it was considered to be addressed later in the review as (conducted after 2010) as previously described (12), otherwise the study was excluded. Each research article was screened for relevant information, recorded in the data extraction file (Microsoft Excel). Moreover, data from each method section was extracted using a predefined set of variables, study characteristics, type of participants, study population size, geographical region, and the study conduction period.

### 2.3 Assessment of quality

Each included article was evaluated based on a framework for making a summary assessment of the quality. The related published literature was crossed, then a framework will be structured specifically to determine the level of representativeness of the studied population and to judge the strength of the estimates provided. Six questions are to be answered in each article, and each answer represents either 1 score for yes, 0 score for No or 0 score for not available; A total score for risk of bias and quality was calculated by adding up the scores in all six domains, resulting in a score of between 0 and 6. The highest score indicates the highest quality. Studies with a score for quality greater or equal to 3 (higher quality) were included in the review as previously described (12).

The six domains were: is the study objective clearly defined? is the study sample completely determined? is the study population clearly defined and specified? is participants’ response rate above 70%, are the methodology and the data analysis used rigorous?.

The Trim and Fill method was used to assess the risk of publication bias in each question response in the included studies (14). Publication bias was evaluated separately for each question-corresponding responses only if the question was addressed and answered in studies equal to or greater than ten.

### 2.4 Quantitative analysis

Meta-analysis was performed - whenever possible using Review Manager Software (Version 5.3). In studies where the Standard Error (SE) is not reported, the following formula was used to calculate it: SE□=□√p (1-p)/ n where p stands for Prevalence. The software automatically provided the Confidence Interval (CI) according to the calculated SE, if the CI is provided in a study; it was introduced accordingly. The heterogeneity of each meta-analyses was assessed. The random effects were favored over the fixed effects model in all meta-analysis established as differences between studies are predicted to be possible due to the diversity of the study populations. Sensitivity analysis was also approached to determine the effect of studies conducted on populations which are proposed to behave in indifferent manners or proposed to be more aware of the overall pooled prevalence. Moreover, subgroup analysis was also conducted-whenever suitable to determine awareness level in a specific country or population. A question to participate in the meta-analysis must be included in at least three studies, and questions with similar outcomes were proposed to be the same.

## 3. Results

### 3.1 Characteristics of the Included Studies

A total of 11,038 published articles were identified from the search strategy. Also, we involved the hand search of reference lists of included original research articles and reviews. Among these articles, 163 articles met our inclusion criteria and passed the quality assessment procedure. Twenty-nine articles have reported the prevalence of microbiological infections, while 34 articles have determined the vaccination status among African HCWs. Furthermore, 108 included studies have assessed the awareness and attitude toward different microbial infections. The oldest studies were published in 2010, while the most recent ones were published in 2020. Figure 1 shows the PRISMA flow diagram. The included articles in this meta-analysis and the assessment of their quality are illustrated in S1 Table and S2 Table, respectively. Three hundred forty-seven questions were summarized, among which 96 questions were analyzed and synthesized. Publication bias assessment indicated no major asymmetry.

**Figure 1.**
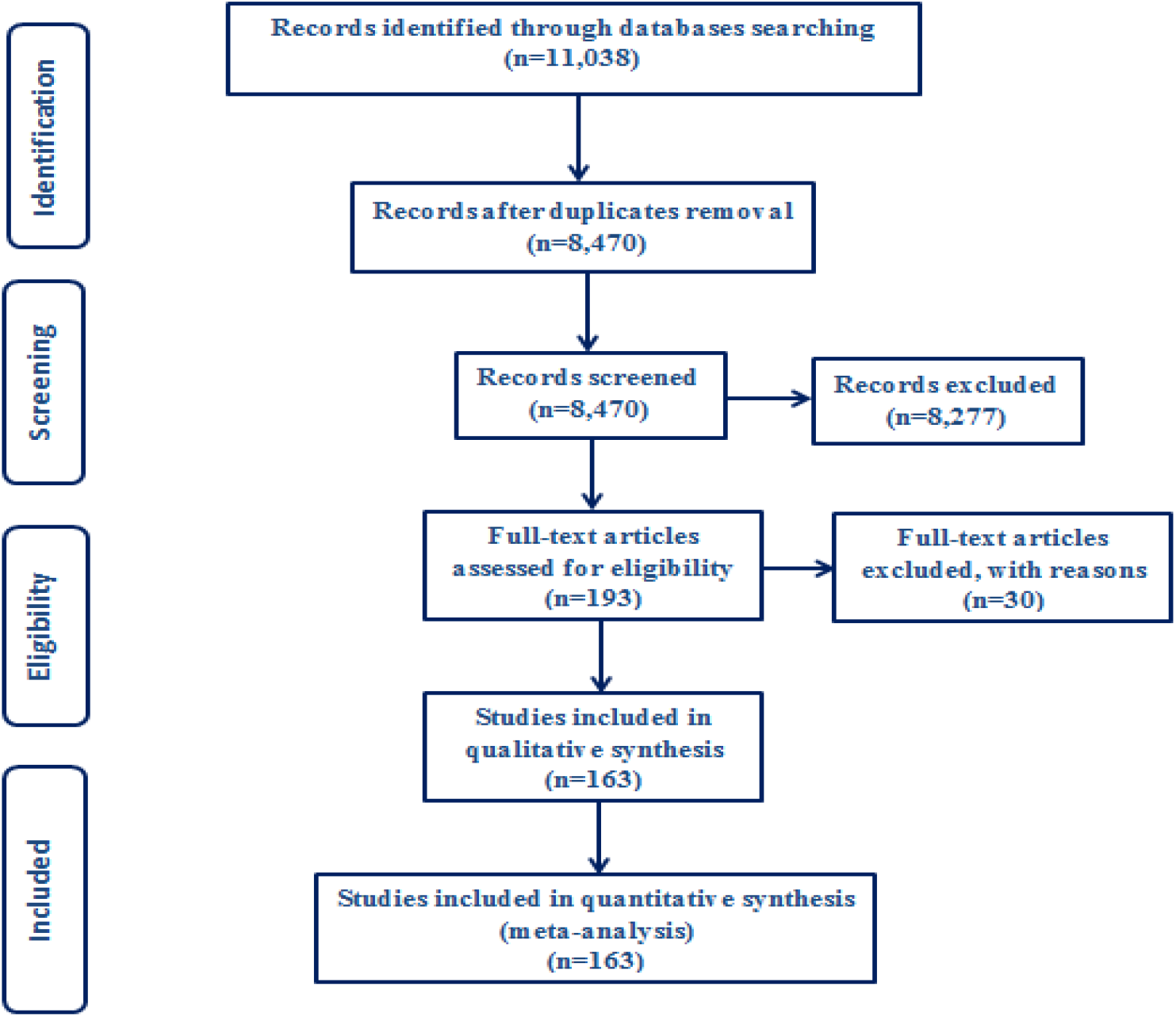
Literature search and selection of studies (PRISMA flow diagram)

### 3.2 Microbiological Infections Among African HCWs

Twenty-nine included studies have assessed the prevalence of different types of microbial infections among 12,342 African HCWs in a total of 12 countries. Eight studies were conducted in Ethiopia (15)(16)(17)(18)(19)(20)(21)(22), four in Cameroon (23)(24)(25)(26), three in each of South Africa (27)(28)(29) and Tanzania (30)(31)(32), two in each of Kenya (33)(34), Sierra Leone (35)(36) and Ghana (37)(38); one in each of Egypt (39), Nigeria (40), Mozambique (41), Sudan (42) and Madagascar (43). The conduction of the studies ranged from 2009 to 2018. Twenty variables were collected from the participants, and 12 of them were analyzed and synthesized. Variables analyzed, their corresponding articles’ data, the pooled prevalence and the confidence intervals are illustrated in Table 1.

**Table 1.** Prevalence of microbiological infections among African HCWs

| Variable | Countries | Total sample size | References | Pooled prevalence | Heterogeneity | Test for overall effect | Type of effect |
| --- | --- | --- | --- | --- | --- | --- | --- |
| HBsAg-positive | Cameroon, Kenya, Sierra Leone, Tanzania, South Africa, Ethiopia, Egypt, Nigeria, Ghana | 6599 | (23)(24)(25)(36)(35)(44)(31)(16)(17)(33)(39)(40)(37)(27) | 6.00 [3.66, 8.33] | Chi<sup>2</sup> = 8.08, df = 13 (P = 0.84); I<sup>2</sup> = 0% | Z = 5.03 (P < 0.00001) | Fixed Effect |
| anti-HCV-positive | Tanzania, Egypt, Ethiopia | 1489 | (23)(39)(17) | 0.81 [-4.27, 5.89] | Chi<sup>2</sup> = 0.02, df = 2 (P = 0.99); I<sup>2</sup> = 0% | Z = 0.31 (P = 0.75) | Fixed Effect |
| HIV-positive | Cameroon, South Africa, Kenya | 1705 | (18)(26)(32) | 9.06 [-6.03, 24.16] | Tau<sup>2</sup> = 157.14; Chi<sup>2</sup> = 18.59, df = 2 (P < 0.0001); I<sup>2</sup> = 89% | Z = 1.18 (P = 0.24) | Random Effect |
| S. aureus (nasal colonization) | Ghana, Sudan, Madagascar | 1093 | (42)(38)(43) | 13.36 [7.87, 18.85] | Chi<sup>2</sup> = 3.66, df = 2 (P = 0.16); I<sup>2</sup> = 45% | Z = 4.77 (P < 0.00001) | Fixed Effect |
| Occupational exposure to blood-borne viruses (HBV, HCV and HIV) | Ethiopia, Tanzania | 1017 | (15)(17)(18)(32) | 81.20 [69.57, 92.82] | Tau<sup>2</sup> = 321.92; Chi<sup>2</sup> = 150.36, df = 4 (P < 0.00001); I<sup>2</sup> = 97% | Z = 8.38 (P < 0.00001) | Random Effect |
| Latent TB (LTB)-positive by tuberculin skin test (TST) | South Africa, Kenya, Mozambique | 1436 | (28)(34)(41) | 59.82 [36.90, 82.75] | Tau<sup>2</sup> = 397.14; Chi<sup>2</sup> = 104.19, df = 2 (P < 0.00001); I<sup>2</sup> = 98% | Z = 5.12 (P < 0.00001) | Random Effect |
| Needle stick injury (once) | Egypt, Ethiopia, Sierra Leone | 1230 | (18)(36)(20)(39) | 14.68 [9.55, 19.80] | Chi<sup>2</sup> = 1.93, df = 3 (P = 0.59); I<sup>2</sup> = 0% | Z = 5.61 (P < 0.00001) | Fixed Effect |
| Needle stick injury (twice) | Egypt, Ethiopia, Sierra Leone | 1035 | (18)(36)(39) | 16.18 [-0.68, 33.04] | Tau<sup>2</sup> = 191.80; Chi<sup>2</sup> = 15.52, df = 2 (P = 0.0004); I<sup>2</sup> = 87% | Z = 1.88 (P = 0.06) | Random Effect |
| History of exposure to needle stick injury (at least once) | Ethiopia, Tanzania | 1830 | (18)(32)(21)(31)(20) | 26.81 [14.58, 39.05] | Tau<sup>2</sup> = 211.99; Chi<sup>2</sup> = 63.30, df = 5 (P < 0.00001); I<sup>2</sup> = 92% | Z = 4.30 (P < 0.0001) | Random Effect |
| History of exposure to blood or body fluids on intact skin | Ethiopia, Tanzania, Guinea | 847 | (15)(32)(45) | 68.57 [38.66, 98.49] | Tau<sup>2</sup> = 684.77; Chi<sup>2</sup> = 115.71, df = 2 (P < 0.00001); I<sup>2</sup> = 98% | Z = 4.49 (P < 0.00001) | Random Effect |
| History of splash of blood or body fluids to eye or mouth | Ethiopia, Tanzania | 1417 | (15)(21)(31) | 64.15 [39.69, 88.61] | Tau<sup>2</sup> = 459.86; Chi<sup>2</sup> = 134.64, df = 2 (P < 0.00001); I<sup>2</sup> = 99% | Z = 5.14 (P < 0.00001) | Random Effect |
| Sharps injury (at least once) for one year | Ethiopia, South Africa | 1307 | (19)(20)(22)(29) | 27.83 [14.29, 41.37] | Tau<sup>2</sup> = 166.53; Chi<sup>2</sup> = 25.24, df = 3 (P < 0.0001); I<sup>2</sup> = 88% | Z = 4.03 (P < 0.0001) | Random Effect |

#### 3.2.1 HBV and HCV

Fourteen studies have investigated the prevalence of HBV, among which three studies have investigated the prevalence of HCV too (44)(17)(39). These studies were conducted in Cameron (23)(24)(25),Sierra Leone (36)(35), Tanzania (44)(31)Ethiopia (16)(17), Kenya (33), Egypt (39), Nigeria (40), Ghana (37) and South Africa (27). The oldest among included studies was conducted in 2012, while the newest was conducted in 2018. The prevalence of HBV (HBsAg) was tested for 6,599 African HCWs;6.00% [95% Cl; 3.66, 8.33] were positive. While the prevalence of HCV (anti-HCV) was investigated for 1,489 African HCWs; 0.81% [95% Cl; −4.27, 5.89] were positive, see Table 1, Figure 2 and Figure 3.

**Figure 2.**
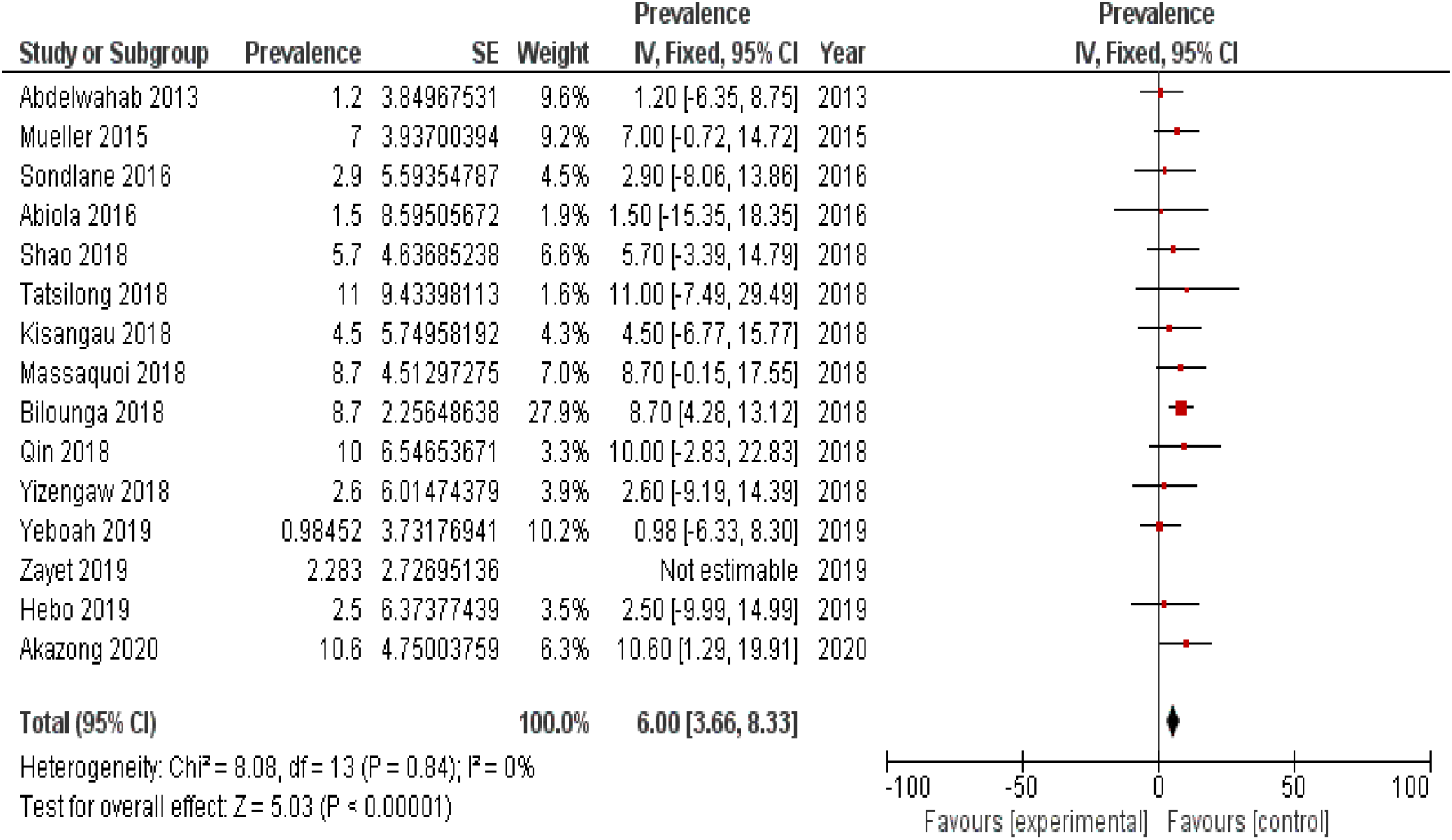
Prevalence of HBV (HBsAg) among African HCWs from studies included in the review.

**Figure 3.**
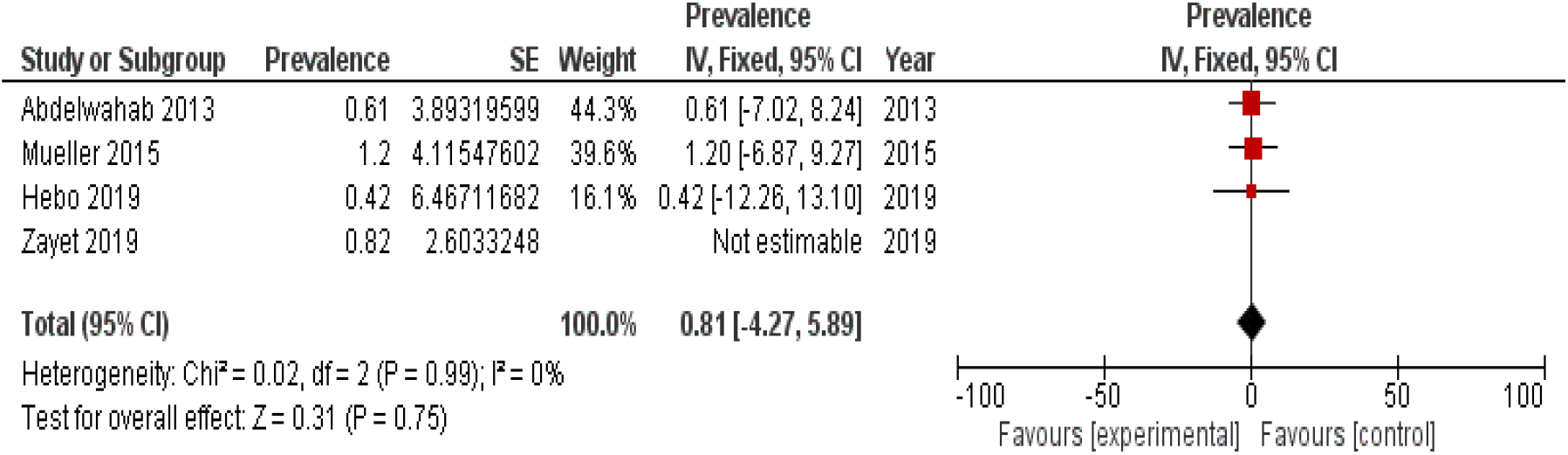
Prevalence of HCV (anti-HCV) among African HCWs from studies included in this review.

#### 3.2.2 Other Microbial Infections

Three articles have studied HIV infection, were conducted in Ethiopia (18), Cameroon (26) and Tanzania (32) in which two studies have investigated the occupational exposure to HIV infection as well (18)(32). In addition, two studies have assessed HIV and TB coinfection which were conducted in South Africa (28) and Kenya (34). The two studies, and another study conducted in Mozambique (41), have also investigated latent tuberculosis infection prevalence (LTB). The prevalence of HIV was assessed for 1,705 African HCWs; 9.06% [95% Cl; −6.03, 24.16] were positive. While the prevalence of occupational Latent TB (LTB) by tuberculin skin test (TST) was 59.82% [95% Cl; 36.90, 82.75] among 1,436 participants, and the heterogeneity was high (I^2^ = 98%).

#### 3.2.3 Occupational Exposure

History of occupational exposure to blood-borne viruses (HBV, HCV and HIV) was assessed for 1,017 African HCWs by four studies conducted in Ethiopia (15)(17)(18) and Tanzania (32). A 81.20% [95% Cl; 69.57, 92.82] were occupationally exposed to blood-borne viruses. Furthermore, three studies have investigated the prevalence of *S. aureus* (nasal colonization) among 1,093 African HCWs;13.36% [95% Cl; 7.87, 18.85] were positive. These studies were conducted in Sudan (42), Ghana (38), Madagascar (43).

Seven studies have assessed the occupational exposure to needle stick injuries, while four have studied the exposure to sharp injuries in general. These studies were conducted in Ethiopia (18)(20)(21)(19)(22), Sierra Leone (36), Tanzania (32)(31), Egypt (39) and South Africa (29). History of exposure to needle stick injuries was investigated for 1,830 participants; 26.81% [95% Cl; 14.58, 39.05] were identified exposed, while the history of exposure to sharp injuries was assessed for 1,307 participants; 27.83% [14.29, 41.37] were found exposed.

Furthermore, a history of exposure to blood or body fluids was assessed by five studies conducted in Ethiopia (15)(21), Tanzania (31)(32) and Guinea (45). Eight hundred and forty-seven African HCWs were asked about their exposure to a splash of blood or body fluid on intact skin; 68.57% [95% Cl; 38.66, 98.49] answered yes. Also, a history of the splash of blood or body fluids to the eye or mouth was investigated by 1,830 participants; 26.81% [95% Cl; 14.58, 39.05] answered yes (Table 1).

### 3.3 Vaccination Status of African HCWs

Thirty-four included studies have assessed the vaccination of 12,785 African HCWs regarding HBV, HIV, influenza, and TB infection in 11 countries. Six studies were conducted in Nigeria (40)(46)(47)(48)(49)(50), five studies in each of Tanzania (30)(51)(52)(44)(31), Ethiopia (15)(53)(20)(54)(55), and Cameroon (56)(23)(24)(57)(58), two studies in each of Ghana (59)(37), Libya (60)(61), Sierra Leone (62)(36), Kenya (33)(63) and South Africa (64)(27), one in each of Senegal (65), Rwanda (66) and Zambia (67). The conduction of the studies ranged from 2009 to 2018 (Table 2). Eight questions were asked to African HCWs related to their vaccination status; however, six questions were analyzed and synthesized. The question “Are you fully vaccinated against HBV?” was answered by 12,036 African HCWs in Tanzania, Nigeria, Ethiopia, Cameroon, Kenya, Ghana, Zambia, South Africa and Libya; 43.22% [95% Cl; 31.22, 55.21] answered yes. The question “Are you unvaccinated against HBV?” were asked to 7,123 HCWs from Tanzania, Nigeria, Ethiopia, Cameroon and Libya; 58.42% [95% Cl; 42.27, 74.57] answered yes. Questions asked, their corresponding articles’ data, the pooled prevalence, value of Heterogeneity, p-value and the confidence intervals are shown in Table 3. Heterogeneity was high in all questions (I^2^ more than 89%), except for the question “Did you take two doses of hepatitis B vaccine?” where I^2^ = 40%.

**Table 2.** Vaccination status of African HCWs

| Questions | Countries | Total sample size | References | Pooled prevalence | Heterogeneity | Test for overall effect | Type of effect |
| --- | --- | --- | --- | --- | --- | --- | --- |
| Did you take only one dose of the hepatitis B vaccine? | Tanzania, Nigeria, Ethiopia, Cameroon, Ghana | 7245 | (51)(40)(53)(23)(30)(49)(50)(37) | 14.99 [5.73, 24.26] | Tau<sup>2</sup> = 153.53; Chi<sup>2</sup> = 68.05, df = 7 (P < 0.00001); I<sup>2</sup> = 90% | Z = 3.17 (P = 0.002) | Random Effect |
| Did you take two doses of the hepatitis B vaccine? | Tanzania, Nigeria, Ethiopia, Cameroon, Ghana | 7245 | (51)(40)(53)(23)(30)(49)(50)(37) | 10.53 [7.74, 13.33] | Chi<sup>2</sup> = 11.72, df = 7 (P = 0.11); I<sup>2</sup> = 40% | Z = 7.39 (P < 0.00001) | Random Effect |
| Did you take the three doses of the hepatitis B vaccine? | Tanzania, Nigeria, Ethiopia, Cameroon, Kenya, Ghana, Zambia, South | 12036 | (51)(40)(53)(23)(30)(49)(15)(33)(59)(63)(57)(67)(47)(48)(27)(24) | 43.22 [31.22, 55.21] | Tau<sup>2</sup> = 677.27; Chi<sup>2</sup> = 1209.45, df = 18 (P < 0.00001); I<sup>2</sup> = | Z = 7.06 (P < 0.00001) | Random Effect |
|  | Africa, Libya |  | (61)(50)(37) |  | 99% |  |  |
| Are you unvaccinated against HBV? | Tanzania, Nigeria, Ethiopia, Cameroon, Libya | 7123 | (51)(40)(53)(23)<br>(15)(52)(46)(57)<br>(31)(20)(61)(50)<br>(37) | 58.42 [42.27, 74.57] | $\text{Tau}^2 = 800.38$; $\text{Chi}^2 = 1401.22$, $\text{df} = 11$ ($P < 0.00001$); $I^2 = 99\%$ | $Z = 7.09$ ($P < 0.00001$) | Random Effect |
| Are you fully vaccinated against HBV? | Tanzania, Ethiopia, Sierra Leone, Libya, Ghana, South Africa, Zambia, Senegal, Nigeria, Cameroon | 4554 | (46)(31)(53)(62)<br>(60)(59)(64)(30)<br>(67)(36)(54)(65)<br>(50)(58) | 41.86 [28.09, 55.64] | $\text{Tau}^2 = 717.30$; $\text{Chi}^2 = 624.37$, $\text{df} = 14$ ($P < 0.00001$); $I^2 = 98\%$ | $Z = 5.96$ ($P < 0.00001$) | Random Effect |
| Did you take any dose of the hepatitis B vaccine? | Ethiopia, Kenya, Cameroon, Zambia, Nigeria, South Africa, Libya | 6849 | (15)(33)(63)(57)<br>(67)(47)(48)<br>(27)(24)(61)(23)<br>(20)(50) | 48.19 [31.77, 64.61] | $\text{Tau}^2 = 815.66$; $\text{Chi}^2 = 1068.09$, $\text{df} = 11$ ($P < 0.00001$); $I^2 = 99\%$ | $Z = 5.75$ ($P < 0.00001$) | Random Effect |

**Table 3.**
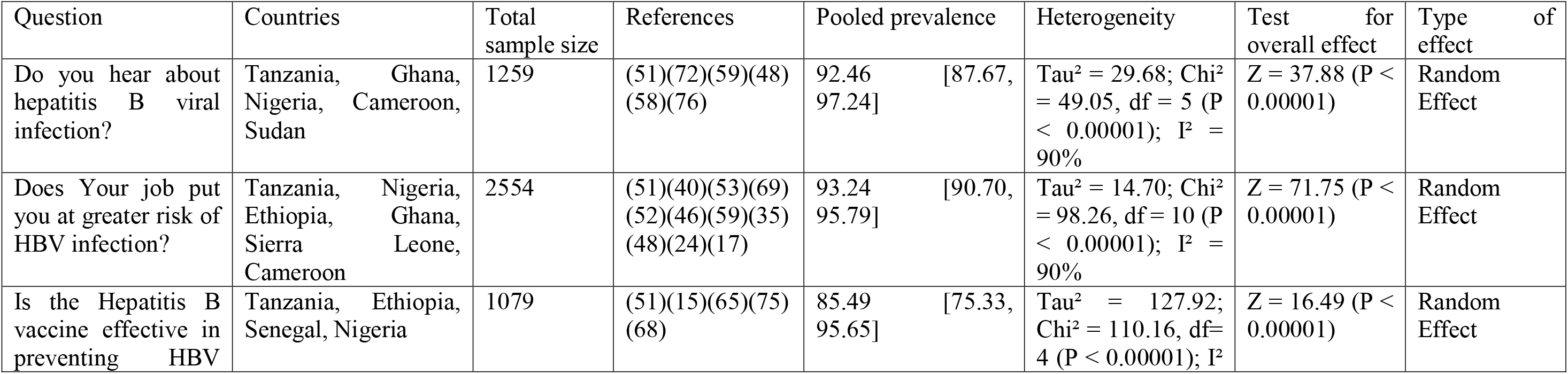
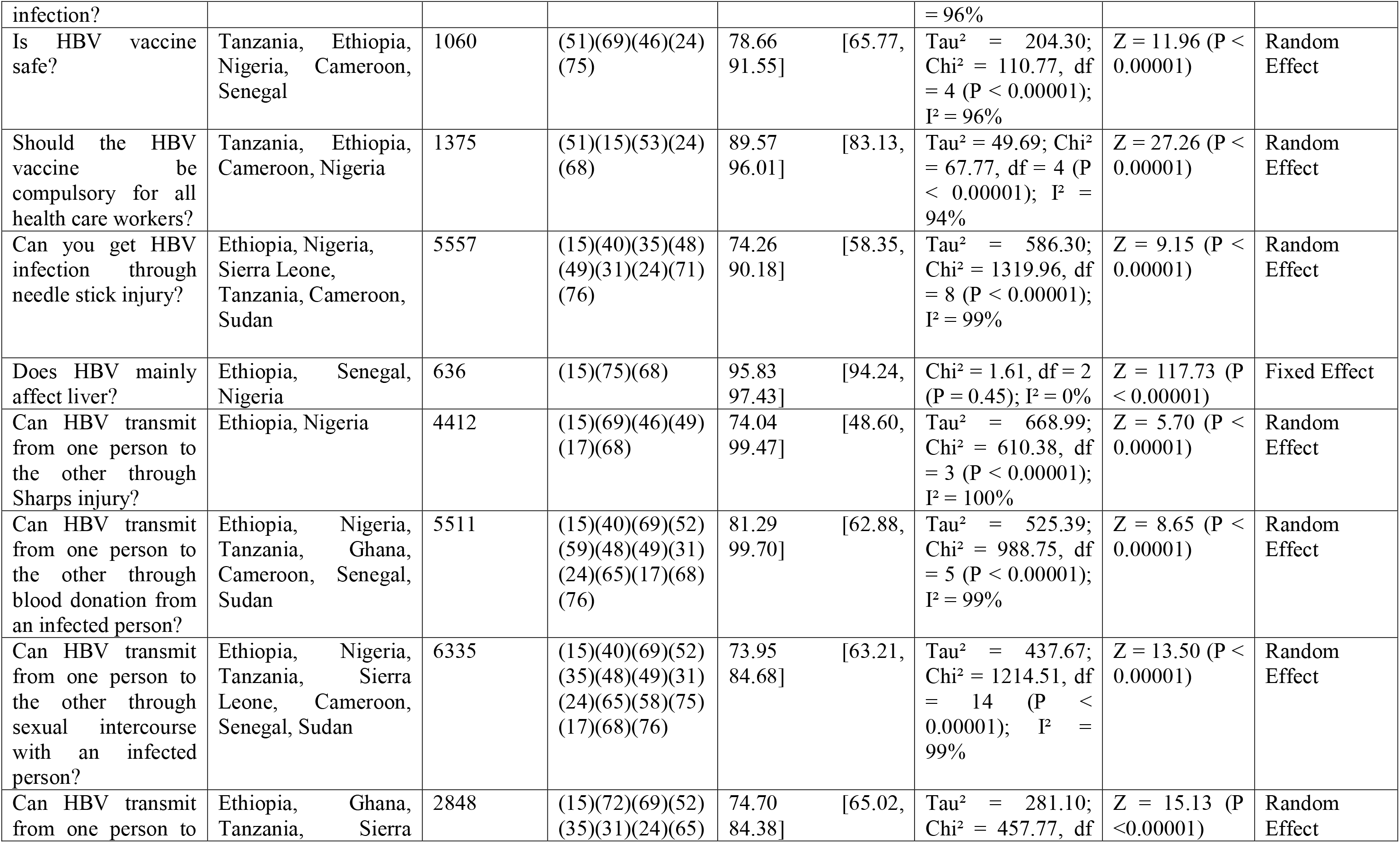
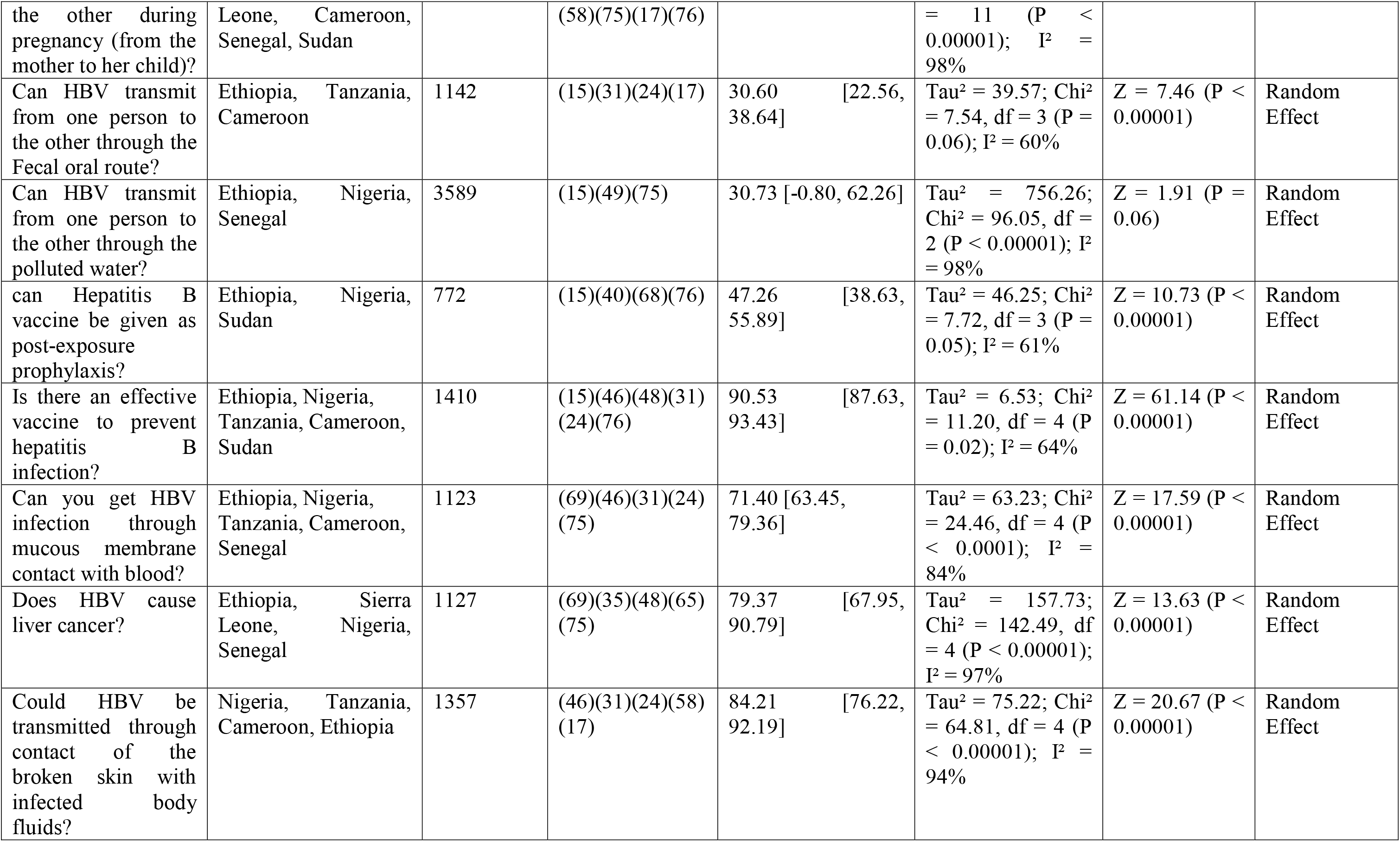
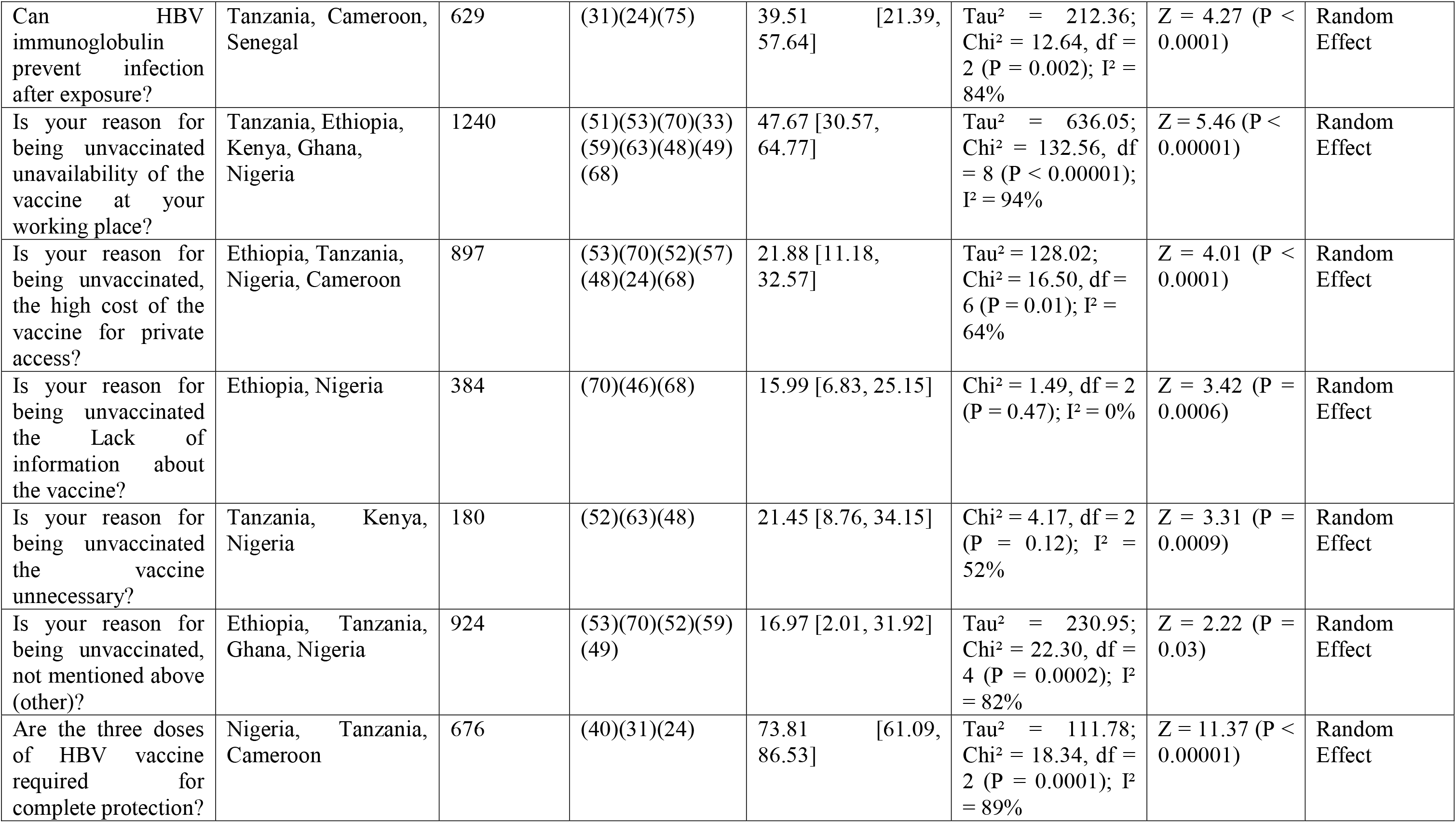
Awareness and attitude toward HBV among African HCWs

### 3.4. Awareness and Attitude among African HCWs

#### 3.4.1 Awareness and attitude toward HBV

Twenty-nine included studies have assessed the awareness and attitude toward HBV among 5,761 African HCWs. These studies were conducted in Tanzania (44)(31)(51)(52), Nigeria (48)(49)(40)(46)(68), Ethiopia (15)(53)(69)(70)(71)(17), Ghana (72)(59), Kenya (33)(63), Côte d’Ivoire (73), Sierra Leone (35)(36), Cameroon (24)(57)(58), Egypt (74), Senegal (65)(75), and Sudan (76). The oldest article among the included studies was conducted in 2010, while the newest was conducted in 2017. Seventy-eight questions related to HBV awareness and attitude were asked to the participants, and 27 of them were analyzed and synthesized. The question “should HBV vaccine be compulsory for all health care workers?” was answered by 1,375Africans HCWs in four countries; 89.57% [95% Cl; 83.13, 96.01] answered yes. While the question “Is your reason for being unvaccinated, unavailability of the vaccine at your working place?” was answered by 1,240 Africans HCWs in five countries; 47.67% [95% Cl; 30.57, 64.77] answered yes. Questions analyzed, their corresponding articles’ data, the pooled prevalence and the confidence intervals are depicted in Table 3.

#### 3.4.2 Awareness and Attitude toward HIV

Nineteen included studies have investigated the awareness of African HCWs regarding HIV; six studies were conducted in Tanzania (77)(78)(32)(79)(80)(81), five in Nigeria (82)(83)(84)(85)(86), four in Ethiopia (18)(71)(55)(22), and one in each of Cameroon (87), Rwanda (66), South Africa (88) and Kenya (89). The oldest among them was conducted in 2010, while the newest was conducted in 2018. Thirty questions were asked to the participants related to their knowledge about HIV, transmission routes, its clinical symptoms, HIV investigation tests, prevention attitude, HIV post exposure prophylaxis (PEP) and their source of information. Among them, 11 questions were analyzed and synthesized (Table 4). The question “Do you consider yourself to be at risk for HIV acquisition at your workplace?” was asked for 1,176; 75.03% [95% Cl; 58.13, 91.93] answered yes. In addition, the question “Ever Heard about HIV Post Exposure Prophylaxis (PEP)?” was asked for 1,013 African HCWs from Nigeria, Cameroon, Tanzania, and Ethiopia; 90.62% [95% Cl; 84.55, 96.69] answered yes. While the question “Ever had training on PEP?” was asked for 494 African HCWs from Cameroon, Ethiopia, and Tanzania; 50.45% [95% Cl; 24.09, 76.81] answered yes. Questions asked, their corresponding study’s countries, the pooled prevalence and the confidence intervals are depicted in Table 4.

**Table 4.** Awareness and attitude toward HIV among African HCWs

| Questions | Countries | Total sample size | References | Pooled prevalence | Heterogeneity | Test for overall effect | Type of effect |
| --- | --- | --- | --- | --- | --- | --- | --- |
| Ever Heard about HIV-Post Exposure Prophylaxis (PEP)? | Nigeria, Cameroon, Tanzania, Ethiopia | 1013 | (83)(87)(80)(84)(55) | 90.62 [84.55, 96.69] | Tau<sup>2</sup> = 41.63; Chi<sup>2</sup> = 93.99, df = 4 (P < 0.00001); I<sup>2</sup> = 96% | Z = 29.25 (P < 0.00001) | Random Effect |
| Are percutaneous injuries considered high-risk exposures for HIV transmission? | Nigeria, Ethiopia | 1334 | (83)(18)(71)(55) | 72.08 [47.80, 96.35] | Tau<sup>2</sup> = 602.09; Chi<sup>2</sup> = 251.54, df = 3 (P < 0.00001); I<sup>2</sup> = 99% | Z = 5.82 (P < 0.00001) | Random Effect |
| Is HIV can transmit through Breast milk? | Nigeria, Cameroon | 360 | (83)(87)(86) | 86.61 [82.83, 90.39] | Chi<sup>2</sup> = 1.84, df = 2 (P = 0.40); I<sup>2</sup> = 0% | Z = 44.91 (P < 0.00001) | Fixed Effect |
| Is HIV can transmit through Saliva? | Nigeria, Cameroon | 357 | (83)(87)(86) | 20.54 [5.37, 35.71] | Tau<sup>2</sup> = 110.41; Chi<sup>2</sup> = 5.20, df = 2 (P = 0.07); I<sup>2</sup> = 62% | Z = 2.65 (P = 0.008) | Random Effect |
| Do you consider yourself to be at risk for HIV acquisition at your workplace? | Nigeria, Cameroon, Ethiopia, Tanzania | 1176 | (83)(87)(18)(79)(55) | 75.03 [58.13, 91.93] | Tau<sup>2</sup> = 360.13; Chi<sup>2</sup> = 244.88, df = 4 (P < 0.00001); I<sup>2</sup> = 98% | Z = 8.70 (P < 0.00001) | Random Effect |
| Is a wash thoroughly with soap and water the initial first-aid measure to institute following needle stick injury? | Nigeria, Cameroon | 407 | (83)(87)(84) | 52.17 [-4.89, 109.23] | Tau<sup>2</sup> = 2501.11; Chi<sup>2</sup> = 159.31, df = 2 (P < 0.00001); I<sup>2</sup> = 99% | Z = 1.79 (P = 0.07) | Random Effect |
| Ever had training on PEP? | Cameroon, Ethiopia, Tanzania | 494 | (87)(18)(80) | 50.45 [24.09, 76.81] | Tau<sup>2</sup> = 496.76; Chi<sup>2</sup> = 34.68, df = 2 (P < 0.00001); I<sup>2</sup> = 94% | Z = 3.75 (P = 0.0002) | Random Effect |
| What is the duration of | Nigeria, Cameroon, | 559 | (83)(87)(55) | 70.23 [50.73, | Tau<sup>2</sup> = 266.43; | Z = 7.06 (P | Random |
| HIV -PEP: 4 weeks? | Ethiopia |  |  | 89.72] | Chi<sup>2</sup> = 48.45, df = 2 (P < 0.00001); I<sup>2</sup> = 96% | <0.00001) | Effect |
| Should HIV -PEP be administered for needle stick injury? | Nigeria, Cameroon, Ethiopia | 227 | (83)(87)(55) | 38.03 [1.34, 74.73] | Tau<sup>2</sup> = 962.82; Chi<sup>2</sup> = 26.18, df = 2 (P < 0.00001); I<sup>2</sup> = 92% | Z = 2.03 (P = 0.04) | Random Effect |
| How soon after needle stick injury should PEP be commenced: within 1 hour | Nigeria, Cameroon, Ethiopia | 555 | (83)(87)(55) | 84.95 [75.43, 94.47] | Tau<sup>2</sup> = 57.91; Chi<sup>2</sup> = 18.93, df = 2 (P < 0.0001); I<sup>2</sup> = 89% | Z = 17.49 (P < 0.00001) | Random Effect |
| How soon after needle stick injury should PEP be commenced: after 72 hours | Nigeria, Cameroon, Ethiopia | 555 | (83)(87)(55) | 10.52 [-2.88, 23.92] | Tau<sup>2</sup> = 84.50; Chi<sup>2</sup> = 5.08, df = 2 (P = 0.08); I<sup>2</sup> = 61% | Z = 1.54 (P = 0.12) | Random Effect |

#### 3.4.3 Awareness and attitude toward TB

Thirteen published articles have studied the awareness and attitude of TB among 2,765 in a total of five countries, four in Ethiopia (90)(91)(54)(92), three in both Nigeria (93)(94)(95) and Lesotho (96)(97)(98), two in South Africa (88)(99) and one in Uganda (100). The conduction of the studies ranged from 2010 to 2015. Sixty-three questions were asked to the participants related to the administrative, environmental and personal tuberculosis control measures, knowledge toward tuberculosis transmission, predisposing factors, prevention, diagnosis and treatment, and attitudes towards preventive methods of nosocomial tuberculosis. However, 14 questions were analyzed and synthesized. The question “Doses HIV consider a predisposition to contracting TB?” were answered by 1,547 participants; 77.11% [95% Cl; 68.48, 85.74] answered yes. Also, the question “Do you agree that surgical masks do not protect the wearer against TB infection?” was answered by 1,316 participants; 35.28 [95% Cl; 30.90, 39.65] answered yes.

Heterogeneity was high in all questions (I^2^ more than 80%). Except for the question “Do you agree that surgical masks do not protect the wearer against TB infection?” where I^2^ = 40%. Questions asked, their corresponding articles’ data, the pooled prevalence, value of Heterogeneity, P-value and the confidence intervals are shown in Table 5.

**Table 5.**
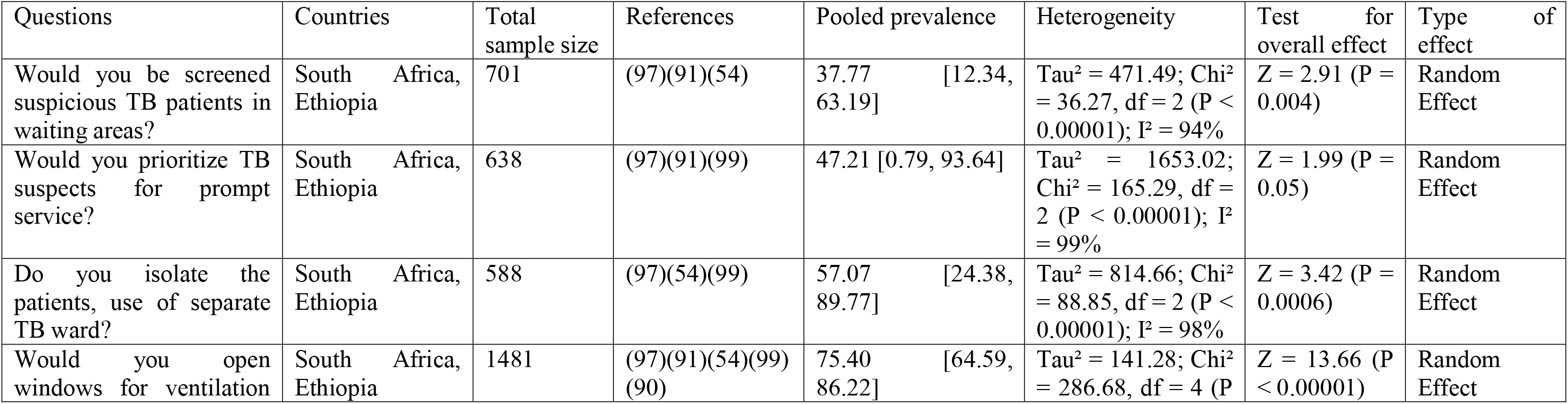
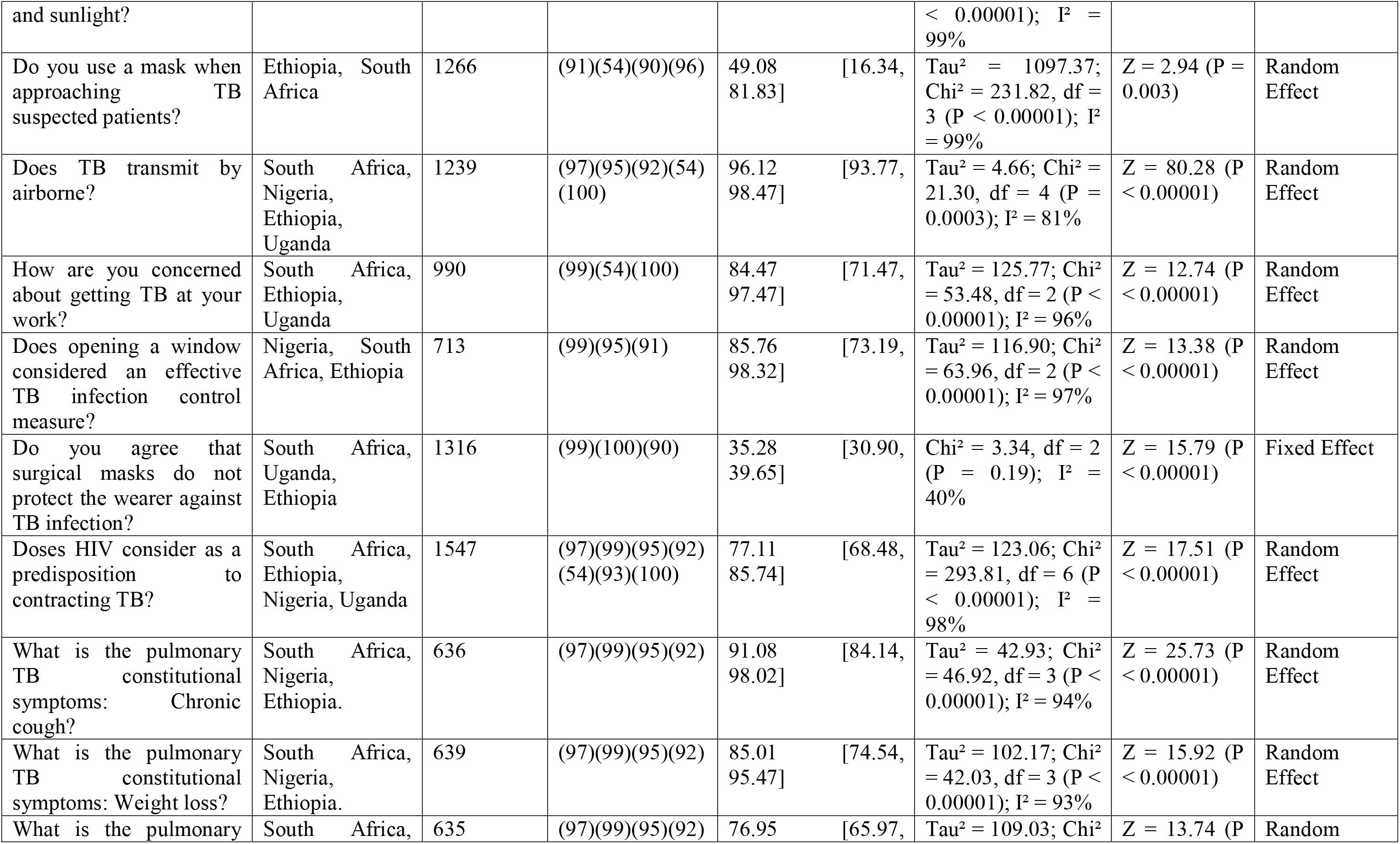
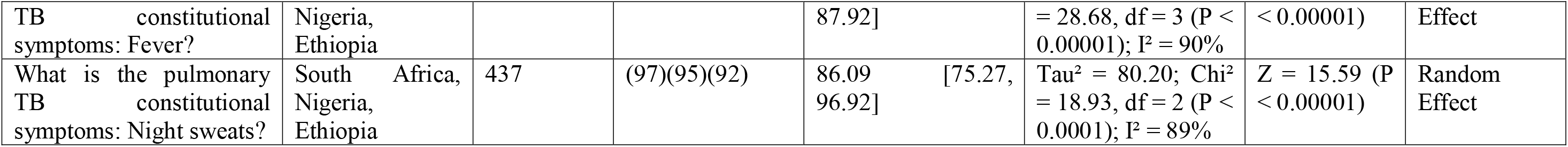
Awareness and attitude toward TB among African HCWs.

#### 3.4.4 Awareness & Attitude toward Antibiotics

Eight studies have assessed the awareness toward antibiotics among 1,761 African HCWs in Ethiopia (101), Gambia (102), Ghanaian (103), Egypt (104), Nigeria (105), Uganda (106) and Lesotho (96). In addition, Bulabula*et al.* performed an online questionnaire (SurveyMonkey) in both French and English, which was circulated via the Infection Control Africa Network (ICAN)/ BSAC on the education and management of antimicrobials amongst nurses from different African countries Democratic Republic of Congo (DRC), Egypt, Kenya, Liberia, Namibia, Nigeria, Rwanda, Sierra Leone, South Africa, Swaziland, Senegal and Zimbabwe (107). The conduction of the studies ranged from 2010 to 2016. Fifteen antibiotics questions were asked, in which three are analyzed and synthesized; see Table 6 for more illustration.

**Table 6.** Awareness and attitude toward antibiotics among African HCWs.

| Questions | Countries | Total sample size | References | Pooled prevalence | Heterogeneity | Test for overall effect | Type of effect |
| --- | --- | --- | --- | --- | --- | --- | --- |
| Does widespread or overuse of antibiotics promote antimicrobial resistance (AMR)? | Ethiopia, Gambia, Ghanaian | 757 | (101)(102)(103) | 77.75 [71.99, 83.52] | Tau<sup>2</sup> = 15.48; Chi<sup>2</sup> = 5.03, df = 2 (P = 0.08); I<sup>2</sup> = 60% | Z = 26.45 (P < 0.00001) | Random Effect |
| Does patients' poor adherence to prescribed antibiotics promote AMR? | Ethiopia, Egypt, Ghanaian | 863 | (101)(104)(103) | 47.18 [-2.80, 97.16] | Tau<sup>2</sup> = 1929.33; Chi<sup>2</sup> = 218.24, df = 2 (P < 0.00001); I<sup>2</sup> = 99% | Z = 1.85 (P = 0.06) | Random Effect |
| As a local factor, does self-prescription by patients develop AMR? | Ethiopia, Egypt, Ghanaian | 858 | (101)(104)(103) | 42.00 [18.79, 65.20] | Tau<sup>2</sup> = 398.18; Chi<sup>2</sup> = 39.55, df = 2 (P < 0.00001); I<sup>2</sup> = 95% | Z = 3.55 (P = 0.0004) | Random Effect |

According to our analysis, the most crucial perceived factors that were determined by African HCWs, contributing to the development of antimicrobial resistance were: widespread or overuse of antibiotics 77.75% [95% Cl; 71.99, 83.52] and patients’ poor adherence to prescribed antibiotics 47.18% [95% Cl; −2.80, 97.16]. Furthermore, the most important local factor identified among respondents for the spread of AMR was self-antibiotic prescription 42.00 % [18.79, 65.20]. Questions asked, their corresponding articles’ data, the pooled prevalence, value of Heterogeneity, P-value and the confidence intervals are shown in Table 6.

#### 3.4.5 Awareness & Attitude toward Infection Control

Twenty-one included studies have investigated the awareness, attitude and practice of infection control measures among African HCWs who were from Nigeria (108)(109)(110)(111)(48)(112)(113), Ethiopia (22)(114)(71)(115)(20)(54)(71)(17), Uganda (116)(117), Egypt (74), Kenya (118), Sierra Leone (119) and Benin (120). The oldest study was conducted in 2010, while the newest was conducted in 2018. Sixty-eight questions were asked to the participants regarding knowledge, attitude, and practice of infection control; however, 18 were analyzed and synthesized. The question “Does the infection prevention and control (IPC) guideline available in your workplace?” was asked to 1,582 HCWs from Ethiopia and Sierra Leone; 50.95% [95% Cl; 40.22, 61.67] answered yes. Regarding the prevention measures against needle stick Injuries, the question “Do you practice recapping syringes?” was asked to 742 HCWs; 43.41% [95% Cl; 23.78, 63.04] answered yes. While, the question “Do you use alcohol swab after needle stick?” was answered by 666 HCWs, and 63.80% [95% Cl; 47.85, 79.74] of them answered yes. In addition, the question “Do you know that the safety box should be closed/sealed when three quarters filled?” was asked to 1,170 HCWs from Ethiopia; 65.80% [95% Cl; 44.09, 87.51] answered yes. Questions asked, their corresponding articles’ data, the pooled prevalence, value of Heterogeneity, P-value and the confidence intervals are depicted in Table 7.

**Table 7.**
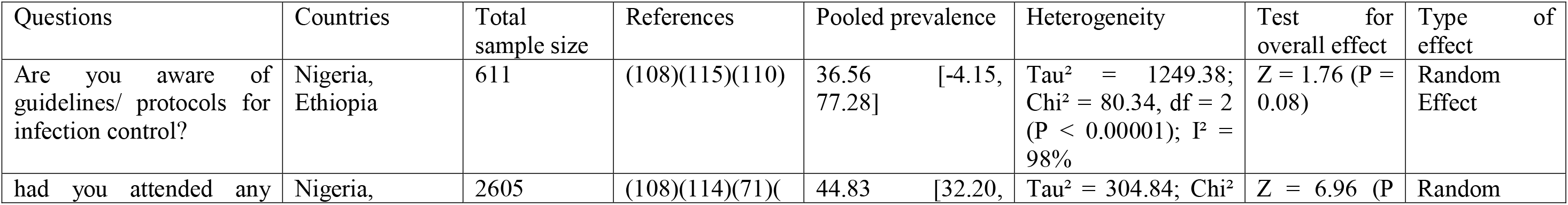
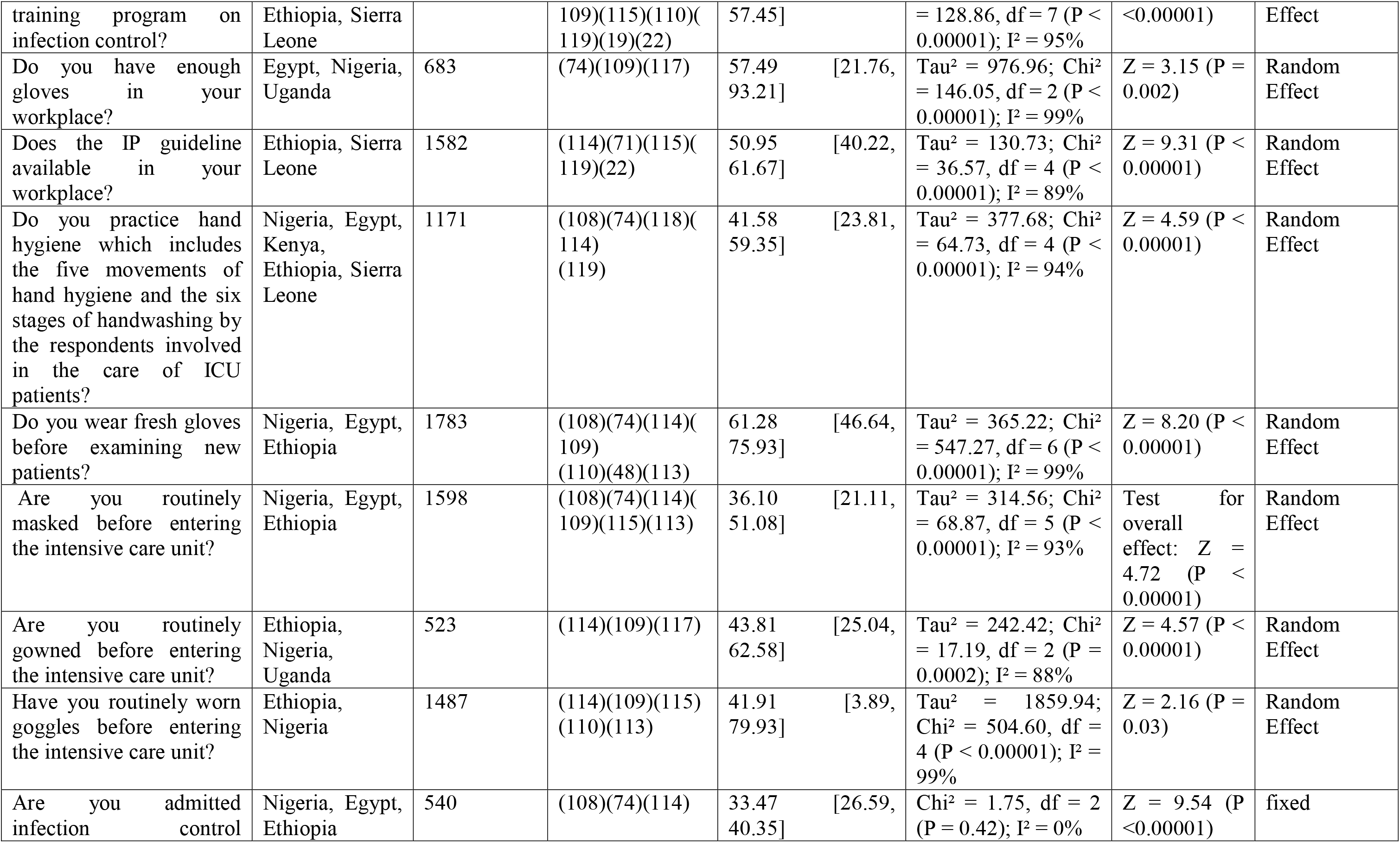
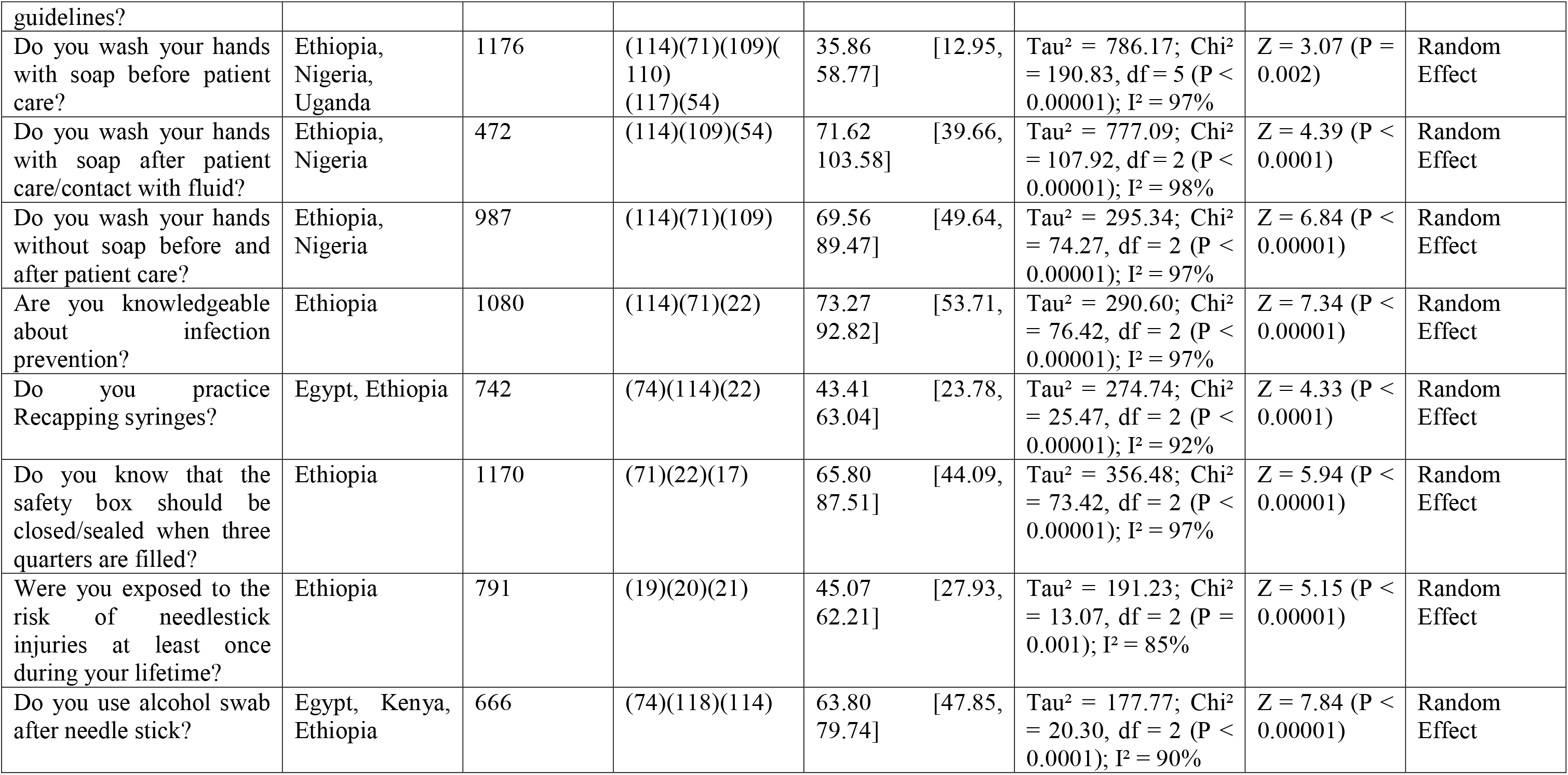
Awareness and attitude toward infection control among African HCWs.

#### 3.4.6 Miscellaneous studies

Eighteen studies have assessed awareness and attitude of African HCWs toward different aspects, including Ebola virus, Influenza virus, Zika virus, zoonotic diseases (rabies, anthrax, dengue, and chikungunya), Buruli ulcer disease, human papillomavirus (HPV), atopic dermatitis, Lassa fever, aflatoxin contamination in groundnut and its ingestion risk, compassion practice towards HIV patients (nursing practice), and Provider-initiated testing and counselling (PITC) for HIV. These articles were from Nigeria (121)(122)(109)(123), Uganda (124)(125), Cote d’Ivoire (126), Cameroon (127), Ethiopia (128), Sierra Leone (62), Tanzania (129)(81) and South Africa (64)(130). Thirty-eight questions were summarized, among which only one question was analyzed and synthesized. The question was, “do you agree that Influenza is more serious than a “common cold”, and patients to be hospitalized or die from influenza?”. It was asked for 1,096 African HCWs from Cote d ’ Ivoire(126), Sierra Leone (62) and South Africa (64), however, 90.50% [95% Cl; 82.85, 98.15] answered yes (Tau² = 42.82; Chi² = 34.47, df = 2 (P < 0.00001); I² = 94%).

## 4. Discussion

This systematic review has attempted, for the first time, to collate all research conducted in Africa reporting any aspect of microbiological assessment among HCWs. The results presented analyzed data about the prevalence of microbial infections, vaccination status and awareness and attitude toward microbial infections among African HCWs to plan and implement adequate preventive measures, optimize practitioners’ knowledge and inform researchers what is done in this area while simultaneously identifying potential gaps in knowledge on this area of study.

The current study has assessed the prevalence of different types and causes of microbial infections among 12,342 African HCWs in 12 countries. The prevalence of HBV (HBsAg), HCV (anti-HCV) and HIV were 6.00%, 0.81% and 9.06%, respectively. The risk of HBV, HCV or HIV infecting HCWs is higher than general populations because they are frequently exposed to potentially infectious biological materials. However, HBV and HCV epidemiology exhibits considerable geographical differences, and accordingly, the prevalence of HBV and HCV among HCWs varies as well (for a review, see (131)). Regarding HIV epidemiology, the Sub-Saharan African belt bears the brunt of HIV infection (132)(133)(134), and HCWs are at high risk of HIV infection from both occupational exposure and sexual transmission (135). Moreover, in this study, the evidence base shows a high burden (59.82%) of occupational Latent TB (LTB) among African HCWs. However, high LTB incidence was also reported in India among HCWs (136)(137). Although HCWs potentially expose to a different type of occupational hazard, many HCWs lack awareness about prevention and control. In African and developing countries, prevention policies are unclear, inaccessible, or attitude problem that made HCWs suffer (138). In this study, 81.20% of African HCWs were occupationally exposed to blood-borne viruses (HBV, HCV, and HIV). The global burden of blood-borne diseases from occupational exposure, based on the world health organization estimation, is around 40% of the hepatitis B and C infections and 2.5% of the HIV infection (139)(140)(141). In addition, occupational exposure to sharp and needlestick injuries represents a global problem. In the USA, According to the CDC report, the annual number of injuries among the hospital staff is 385,000 (142). While in Europe, over 1 million needle-stick injuries are suffered by healthcare workers each year (143). Moreover, the reported incidence rates of needlestick injuries annually were 100/1000 healthcare workers in Finland and 2.2/100 full-time equivalent physicians and 7.0/100 nurses in France (144)(145). In this study, the pooled prevalence of historical exposure to needle stick injuries and sharp injuries were 26.81% and 27.83% among 1,830 and 1,307 African HCWs, respectively.

Being a healthcare worker put you endanger of acquiring a vaccine-preventable disease (VPD) (146). As for this, vaccination was a very effective and safe way to prevent VPD (147). According to WHO, there are vaccines available to protect against at least 20 diseases (147), amongst which HBV vaccine, which is considered the most important one for HCWs providing the fact that hepatitis B infection is four times greater than that of the general adult population (148). Despite this, in our study, we found that 58.42% (n= 7,123) of HCWs in Africa were unvaccinated in comparison to 46.1% in India (149), 24% in Turkey (150) and 19.3% in Brazil (148). In contrast, vaccination coverage was 43.22% (n= 12,036) which is lower when compared to 77% in each of Italy, Australia and New Zealand (151), China 60% (152), Pakistan 73.42% (153), India 56% (154), Iran 48.1% (155) and USA 63.4%, and is the highest in French healthcare students (91.8%) probably for mandatory vaccination (156)(157). Therefore, because of excellent vaccination coverage in France and compulsory vaccination and given that high hepatitis B infection rate among African HCWs offset by low vaccination coverage, we strongly recommend HBV vaccine to be an obligatory prerequisite for work. This policy will greatly value HCWs regarding their health state and, therefore their service.

Measuring awareness toward blood born infections in HCWs is vital since their a high risk of contracting them (158) as the incidence of needle stick injuries are high in their work field (147). Opening with HIV as it demonstrated the highest prevalence among African HCWs, our study found that most of them consider themselves at risk for HIV acquisition at their workplace by 75.03%, which is lower when compared to South India (100%) (158) and Serbia (89%) (3), but higher than the 62% recorded among HCWs in Israel (4). The majority of HCWs have Heard about HIV Post Exposure Prophylaxis (PEP) (90.62%); similarly, high rates were identified in South India (158) and so much less in Serbia (3). Surprisingly only 50.45% had training on PEP in contrast to 68.5% in South India (158). Nevertheless, a much lower rate was reported in Serbia (3). Regarding attitude towards HBV and its vaccination, it was interestingly high the percentage of HCWs stating that vaccines should be compulsory (89.57%) in line with a study done in Iraq (161). Unfortunately, 47.67% of African health care workers declared that the reason for not being vaccinated was the unavailability of the vaccine, which was also mentioned as one of the top reasons in several studies conducted in China (152), Iran (155) and Pakistan (153)(162). Considering this high percentage, the significance of providing the HBV vaccine at work would help in improving the vaccination status among HCWs.

Furthermore, in light of the high burden of TB among African HCWs, knowledge and implementation of TB control measures by HCWs and their compliance and willingness are required to manage healthcare-associated TB effectively. The WHO and CDC proposed numerous comprehensive IPC guidelines which have been designed for healthcare facilities, inclusive of low-resourced settings (163)(164)(165). IPC measures have been proposed at three levels: administrative, environmental and personal respiratory protection. The managerial control measures represent the priority, while the other two are dependent on administrative control measures for their effectiveness (166). In this study, 37.77% screened suspicious TB patients in waiting areas, and 47.21% prioritized TB suspects for prompt service. These poor practices of administrative IPC measures are many pitfalls since these should be implemented at first contact with an infectious patient at a health facility. Delays in screening, diagnosis and treatment of TB would increase the risk of healthcare-associated TB. Environmental IPC measures, which are second-line IPC practices, are dependent on the use of ventilation and irradiation and infrastructural design of the facility; however, more straightforward effective methods have been suggested based on adequate ventilation through the opening of windows (164). in this study, 57.07% of respondents isolate the patients or use of separate TB ward, and 75.40% of respondents open windows for ventilation and sunlight. In addition, the findings regarding PPE were of concern. We found that 49.08% of respondents use a mask when approaching TB suspected patients, which agrees with Biscotto *et al.* finding that HCWs infrequently used masks (39.5%) when performing procedures or attending patients with a high risk of healthcare-associated TB (167).

Antimicrobial resistance (AMR) represents a serious growing challenge, especially in low-income countries such as African countries, because of irrational uses of antimicrobials, lack of clinical microbiology laboratories for antimicrobial susceptibility testing, over-the-counter availability of antibiotics high prevalence of infection (168). However, containment of AMR demands changes in HCWs’ behavior towards the magnitude of AMR problem, prevention the transmission of resistant microbes, prescribing antimicrobials wisely and promoting awareness on AMR for patients and communities (101). Therefore, information on HCWs’ knowledge and awareness of AMR will permit more effective interventions on AMR containment. In this study, the most crucial perceived factors that were determined by African HCWs, contributing to AMR development were widespread or overuse of antibiotics (77.75% by respondents) and patients’ poor adherence to prescribed antibiotics (47.18%). Likewise, previous studies conducted in Scotland, France and Spain found these factors among the leading causes of AMR development (169)(170). In addition, this study revealed that the most important local factor for the development and spread of AMR was self-antibiotic prescription (42% by respondents) which is similar to previous studies (171)(168).

Adherence to IPC guidelines is a key to protect HCWs and prevent transmission of infections, and it becomes even more important when infectious diseases become widespread, such as during the COVID-19 pandemic. In this review, we found that the availability of IPC guidelines in African HCWs’ workplaces was 50.95% which is consistent with a study conducted in Italy (172). Strategies in IPC guidelines include stricter cleaning routines, the separation of patients with respiratory infections from others, and personal protective equipment (PPE) such as masks, gloves and gowns. However, practicing these strategies can be difficult and time-consuming; therefore, authorities and healthcare facilities need to consider the best way to support healthcare workers to implement them. The findings presented in this study regarding the implementation of PPE among African HCWs were 35.86% wash their hands with soap before patient care; 71.62% wash their hands with soap after patient care/contact with fluid; 69.56% wash their hands without soap before and after patient care;61.28% wear fresh gloves before examining new patients; 36.10% are routinely masked before entering the ICU; 43.81% are you routinely gowned before entering the ICU, and 41.91% are routinely worn goggle before entering the ICU (Table 7). These findings are relatively lower compared with results of other similar previous work (173).

## Conclusion

Healthcare workers are god’s safeguard of life, especially in continents suffering poverty, conflicts, illiteracy, and fragile infrastructures. The current study determined many weaknesses to be addressed for the sake of improving health in Africa. Therefore, the current pooled data are critically significant to be used in planning governmental or NGOs strategies.

## Data Availability

All data produced in the present work are contained in the manuscript

## Ethical approval

Not applicable

## Consent for publication

Not applicable

## Data Availability Statement

All relevant data are available within the manuscript and as supplementary materials.

## Funding

The authors received no specific funding for this work.

## Conflict of interest

The authors have declared that no competing interests exist.

## Authors’ contributions

Conceptualization: Marwan M. Badawi

Data curation: Abeer B. Idris, Alaa B. Idris

Data analysis: Elfatih A. Hasabo

Supervision: Marwan M. Badawi

Writing – original draft: Abeer B. Idris, Alaa B. Idris, Marwan M. Badawi, Elfatih A. Hasabo

Writing – review & editing: Nazar Beirag, Abeer B. Idris, Alaa B. Idris, Marwan M. Badawi

## Supplementary files

S1 Table. Characteristics of the included studies.

S1 Table. Assessment of the quality of the included studies.

## Notes

### Competing Interest Statement

The authors have declared no competing interest.

### Funding Statement

This study did not receive any funding

